# Uptake of early infant diagnosis and factors associated with its timely completion among HIV exposed infants at Lira Regional Referral Hospital: a retrospective cohort study

**DOI:** 10.64898/2026.02.28.26347306

**Authors:** Rebecca Awili, Joan Kalyango, Sean Steven Puleh, Joy Acen, Douglas Bulafu, Shaban Rajab Wilobo, Nathan Ntenkaire, Victor Musiime, Eve Nakabembe

## Abstract

**Background:** HIV exposed infants (HEIs) are at a higher risk of infant mortality compared to their counterparts who are not HIV exposed. Early Infant Diagnosis (EID) is the critical first step in reducing HIV-related infant mortality through prompt identification of HIV-infected infants and subsequent initiation of antiretroviral therapy. However, there is limited information on Uptake of EID and factors associated with its timely completion among HIV exposed infants. Therefore, this study aimed at determining the uptake of EID and factors associated with its timely completion among HIV exposed infants at Lira Regional Referral Hospital (LRRH).

**Methods:** The study was a retrospective cohort of 252 HEIs born in the period of 1^st^ January 2021 to 31^st^ December 2021 chosen through consecutive sampling. Data abstraction tools were used to collect data on uptake of 1^st^, 2^nd^, 3^rd^ DNA-PCR and final rapid test from mother-baby pair files and EID register. The main outcome was Uptake of EID and classified as timely and untimely according to the PMTCT guideline. Data was analyzed using descriptive statistics and generalized estimating equations (GEE) with poisson family, log link and unstructured correlation structure.

**Results:** The timely uptake of EID among HIV exposed infants at 4-6 weeks, 9 months, 6 weeks after cessation of breastfeeding and 18 months were 80.1% (95% CI:74.5-84.7), 84.2% (95% CI:79.0-88.3), 3.7% (95% CI:2.0-7.0) and 78.8% (95% CI:73.2-83.6) respectively. Having cotrimoxazole given was associated with timely completion of EID [aRR=2.974, 95% CI (1.45-6.10)]

**Conclusion:** Uptake of EID among HEIs was sub-optimal, below the Ministry of Health’s 90% target. Timely cotrimoxazole administration was associated with EID completion,

## Introduction

Globally, children account for 3 percent of all people living with HIV, 9 percent of new HIV infections and 12 percent of all AIDS-related deaths in 2024(1). Of the estimated 1.38 million children under 15 years of age living with HIV, 86 per cent live in sub-saharan Africa.(1) Sub-Saharan Africa bears the largest burden of pediatric HIV, with about 435 deaths per 100,000 (2). In Uganda, the number of new HIV infection due to Mother-to-child-transmission (MTCT) among children aged 0-14 years was 4,700 in 2024(3).

MTCT accounts for 15-25% of the transmissions that occur during pregnancy and delivery, with an additional 5-20% risk from breastfeeding(4). Interventions like the ‘Test and Treat’ strategy aim to increase early antiretroviral therapy (ART) initiation, particularly through EID for HEIs (5, 6). According to Prevention of Mother to Child transmission (PMTCT) cascade guidelines 2020, HEIs must undergo DNA-PCR testing at 4-6 weeks, 9 months, 6 weeks after cessation of breastfeeding, with a final rapid test at 18 months if prior tests are negative (7).

Despite the availability of these guidelines, EID coverage remains low, with only 64% of HEIs receiving their 1st DNA-PCR test, fewer than 55% receiving virological testing within 2 months, and only 28% completing the final rapid test at 18 months according to a cross sectional study done in Uganda (8). This can be attributed to barriers including; inadequate staffing, late referrals, stigma surrounding HIV infection, lack of social support, HIV disclosure status, and distance to healthcare facility.

A study done in Uganda at Lira Regional Referral Hospital (LRRH) revealed a 51% loss to follow up of mother-infant pairs, with a particularly high loss of 60% between the 2nd PCR and final rapid test, leaving the final HIV status of many infants unknown (9). While this study focused on loss to follow-up, gaps remain in understanding the uptake of DNA-PCR tests and rapid diagnostic tests (RDTs) across all stages as well as the associated factors with timely completion.

A study in Ethiopia has shown antenatal care attendance and maternal counseling on feeding options are associated with timely testing, while poor maternal adherence is linked to higher infants HIV positivity at the 18^th^ month of antibody test (10). However, most studies focus on 1^st^ DNA-PCR, leaving gaps in knowledge about the later stages. Therefore, this study aimed to determine the uptake of EID and factors associated with its timely completion among HEIs at LRRH, providing insights for policymakers to improve EID timely completion.

## Materials and methods

### Study design, period, area and population

This retrospective cohort study utilized data from mother-baby pair files and EID register at EID clinic to assess the uptake of the 1^st^, 2^nd^, 3^rd^ DNA-PCR as well as the final rapid test among HEIs born in the period of 1^st^ January 2021 to 31^st^ December 2021. LRRH is located in Lira city, in Northern Uganda. It’s a referral hospital for districts of Oyam, Kole, Kwania, Apac, Dokolo, Amolatar, Otuke, Alebtong and Lira districts. It’s located about 234 kilometers from Kampala the capital city of Uganda. Funded by the Uganda Ministry of Health, LRRH provides free general and specialized care and has a bed capacity of approximately 400. The hospital’s EID clinic managed around 322 mother-baby pairs for infants born during the study period.

The study included all EID records of HIV-exposed infants born at LRRH within the specified period who met the eligibility criteria. Records were excluded if mother-baby files were missing the infant’s card or if the infants had been transferred out to other health facilities.

### Sample size estimation

Objective 2; assessing factors associated with timely completion of 1^st^, 2^nd^ and 3^rd^ DNA-PCR testing for HIV exposed infants.

Using sample size estimation for proportions in two groups

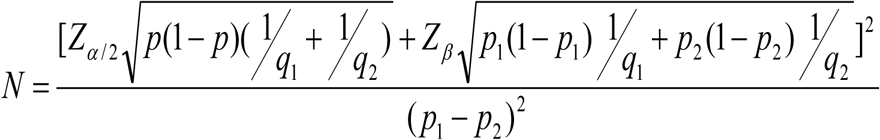

Considering marital status as a factor;

N is the required sample size

q1 is the proportion of subjects in group one, women that are married=0.5

q2 is the proportion of subjects in group two ie women that are not married=0.5

Zα/2-standard normal corresponding with a 5% level of significance=1.96

A zβ-standard normal value corresponding with 80% power=0.84

p1 is the proportion of married women with the outcome of interest ie infants of married participants who adhered to the 1^st^ PCR test =41.1% (11).

p2 is the proportion of unmarried women with the outcome of interest ie infants of single participants who adhered to the 1^st^ PCR test =24.1% (11).

p=p1q1+p2q2=0.326

N=236

Adjusting for missing data of 10%

Estimated sample size=260

Taking the largest, therefore sample size is 260.

### Sampling methods

Consecutive sampling was used to select HIV exposed infants from EID records at LRRH for infants born between January to December 2021.

### Study variables

#### Dependent variables

EID uptake was categorized as timely uptake of 1^st^, 2^nd^, 3^rd^ DNA-PCR and final rapid test according to the timeframe in the WHO guideline 2020 and s uptake when tested after the specified timeframe in the WHO guideline 2020 or not tested

#### Independent variables

Child related characteristics included age, sex, infant feeding, birthweight and infants at risk, maternal related characteristics included age, marital status, time of HIV diagnosis, time to treatment initiation, mode of delivery and most recent viral load, household related characteristics included distance from the health facility and institutional related characteristics included Referrals, provision of ART prophylaxis, entry point

### Data collection methods

The data abstraction form was used to extract information from mother-baby pair files and EID register at EID clinic in LRRH from 3^rd^ July 2023 to 17^th^ July 2023. The data abstraction tool consisted of five sections. The first section collected information on child-related, maternal-related, household-related and institutional-related characteristics. The second, third, and fourth sections were designed to gather data on the 1st, 2nd, and 3rd DNA-PCR tests, respectively. The fifth section captured information on the final rapid test. The abstraction tool was pretested at Ober Health Centre IV using the records of 10 participants to ensure its effectiveness and reliability. Data collection was conducted by the principal investigator (PI) with the assistance of a trained research assistant, a registered midwife.

A two-day training session was held for the data abstraction process and for the research assistant to familiarize herself with the data abstraction process and ensure consistency and accuracy during data collection.

### Data management and analysis

In order to describe baseline socio-demographic and clinical characteristics of participants continuous variables were summarized as means with corresponding standard deviation since data was normally distributed. Categorical variables were summarized as frequencies and percentages. EID uptake was calculated as a percentage of the infants who tested at the four different time points. This was measured at 4-6 weeks, 9 months, 6 weeks after cessation of breastfeeding and 18 months of age which is recommended guideline 2020 along with PMTCT. At bivariate analysis, generalized estimating equations (GEE) with Poisson family, log link and unstructured correlation structure were used to assess the association between the variables and timely completion of EID among HEIs over time. Interaction terms between time and different independent variables were generated and overall chi-square values and related p-values for these interactions were determined. The variables whose interactions with time had p-values <0.2 were considered for multivariate analysis.

At multivariate analysis, variable-time interactions were assessed for statistical significance. In addition, interaction between independent variables was assessed using product terms of variables. Backward elimination of interaction terms was used to drop non-significant interaction terms. Subsequently, confounding was assessed by comparing crude and adjusted relative risks. If a variable changed the relative risk of another by 10% or more, that variable was considered a confounder. Statistical significance in the final model was based on p-value <0.05.

### Quality control measures

A pretest study was conducted at EID clinic at Ober Health Centre IV on 10 participant records. The research assistant underwent a two-day training by the study PI on study protocol and data collection procedures. Double data entry was done and PI continually cross-checked data collection tools for completeness. The PI also continually reviewed performance of the research assistant and held regular meetings.

### Ethical considerations

The study was conducted in accordance with the Helsinki declaration for research involving human subjects. Ethical approval was sought from School of Medicine Research Ethics Committee (Mak-SOMREC-2023-575) of Makerere University College of Health Sciences and administrative clearance was obtained from hospital director, LRRH. A waiver of informed consent was obtained from SOMREC. Data was kept confidential and unique identification numbers assigned to the participants.

## Results, Discussion and Conclusion

### Results

#### Socio-demographic and clinical characteristics of the participants at baseline

The majority of the 252 infants were male (51.2%, n=129). Most of their mothers were married (71.4%, n=180), with a mean age of 32 years, and they came from a distance less or equal to 5 Kilometers (65.9%, n=166). Majority (96%, n=242) of the infants had received ART prophylaxis with 92.9% (n=234) got cotrimoxazole. Socio-demographics and clinical characteristics are summarized in Table 1.

**Table 1.** Socio-demographic and clinical characteristics among 252 HIV exposed infants at LRRH.

| Variable | Category | Mean (SD) | n (%) |
| --- | --- | --- | --- |
| Infant Sex | Male |  | 129 (51.2) |
|  | Female |  | 123(48.8) |
| Marital Status | Married |  | 180(71.4) |
|  | Single |  | 72(28.6) |
| Distance from Health facility | < =5Km |  | 166(65.8) |
|  | >5Km |  | 86(34.2) |
| Mother's age |  | 32.5(6.1) |  |
| Cotrimoxazole given | Yes |  | 234(92.9) |
|  | No |  | 18(7.1) |
| ART Prophylaxis | Yes |  | 242(96.0) |
|  | No |  | 10(4.0) |

*n- Number of participants, % - Percentage, SD-standard deviation.*

#### Uptake of EID at different time points for the 252 HIV-exposed infants from LRRH

The uptake of EID among HIV exposed infants at 4-6 weeks, 9 months, 6 weeks after cessation of breastfeeding and 18 months were 80.1% (95% CI:74.5-84.7), 84.2% (95% CI:79.0-88.3), 3.7% (95% CI:2.0-7.0) and 78.8% (95% CI:73.2-83.6) respectively (Table 2).

**Table 2.**
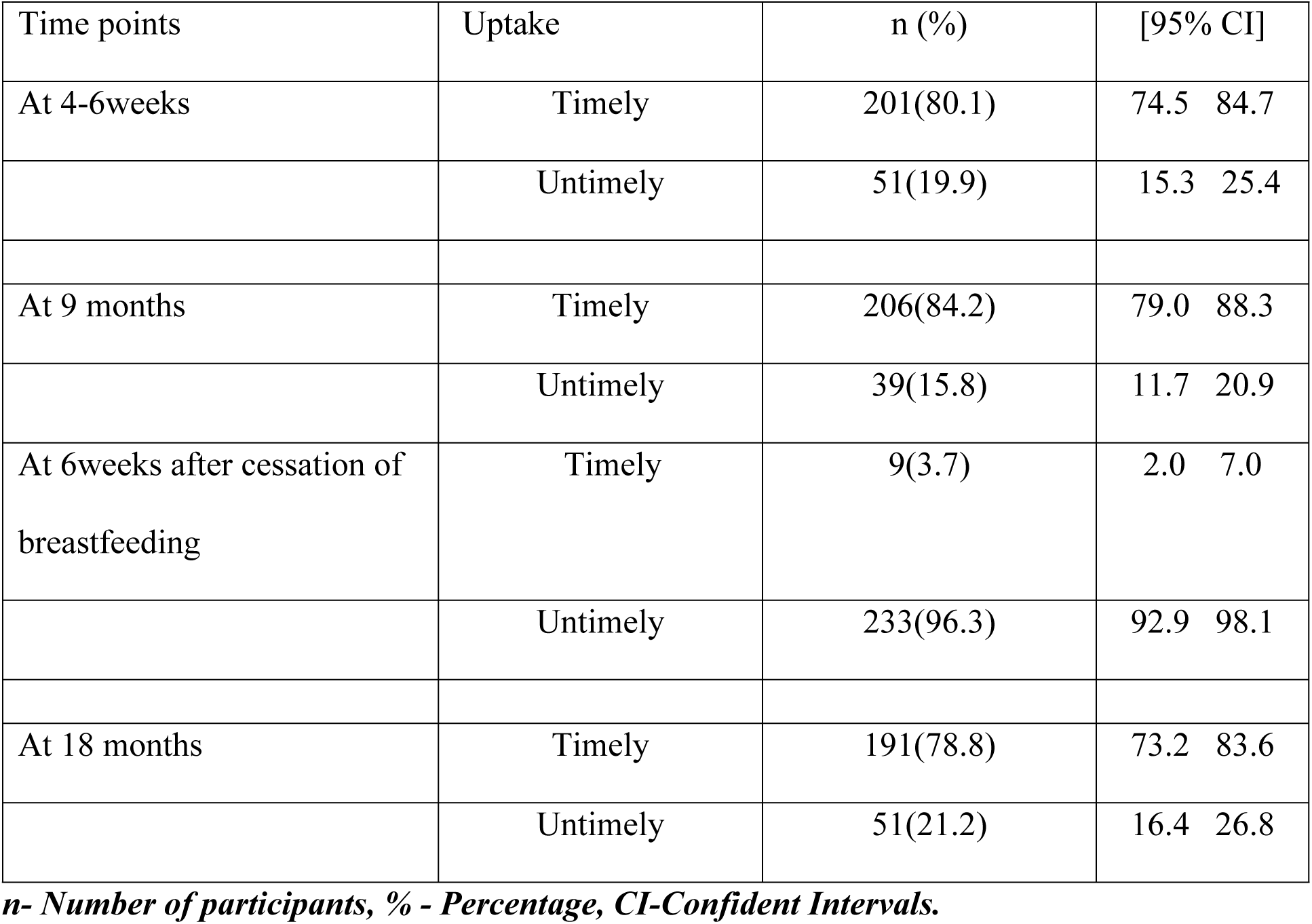
Uptake of EID among HIV exposed infants at LRRH at 4-6 weeks, 9 months, 6 weeks after cessation of breastfeeding and 18 months.

| Time points | Uptake | n (%) | [95% CI] |
| --- | --- | --- | --- |
| At 4-6weeks | Timely | 201(80.1) | 74.5 84.7 |
|  | Untimely | 51(19.9) | 15.3 25.4 |
| At 9 months | Timely | 206(84.2) | 79.0 88.3 |
|  | Untimely | 39(15.8) | 11.7 20.9 |
| At 6weeks after cessation of breastfeeding | Timely | 9(3.7) | 2.0 7.0 |
|  | Untimely | 233(96.3) | 92.9 98.1 |
| At 18 months | Timely | 191(78.8) | 73.2 83.6 |
|  | Untimely | 51(21.2) | 16.4 26.8 |

*n- Number of participants, % - Percentage, CI-Confident Intervals.*

#### Bivariate analysis for factors associated with the timely completion of EID among HIV exposed infants at LRRH

At bivariate analysis, time interaction with infant at risk, cotrimoxazole, mother’s age, time of HIV diagnosis, distance from the health facility, ART prophylaxis and feeding status were had p-values less than 0.2 and were thus considered for multivariate analysis. Results of bivariate analysis for factors associated with timely completion of EID among HIV exposed infants over time are summarized in Table 3

**Table 3.**
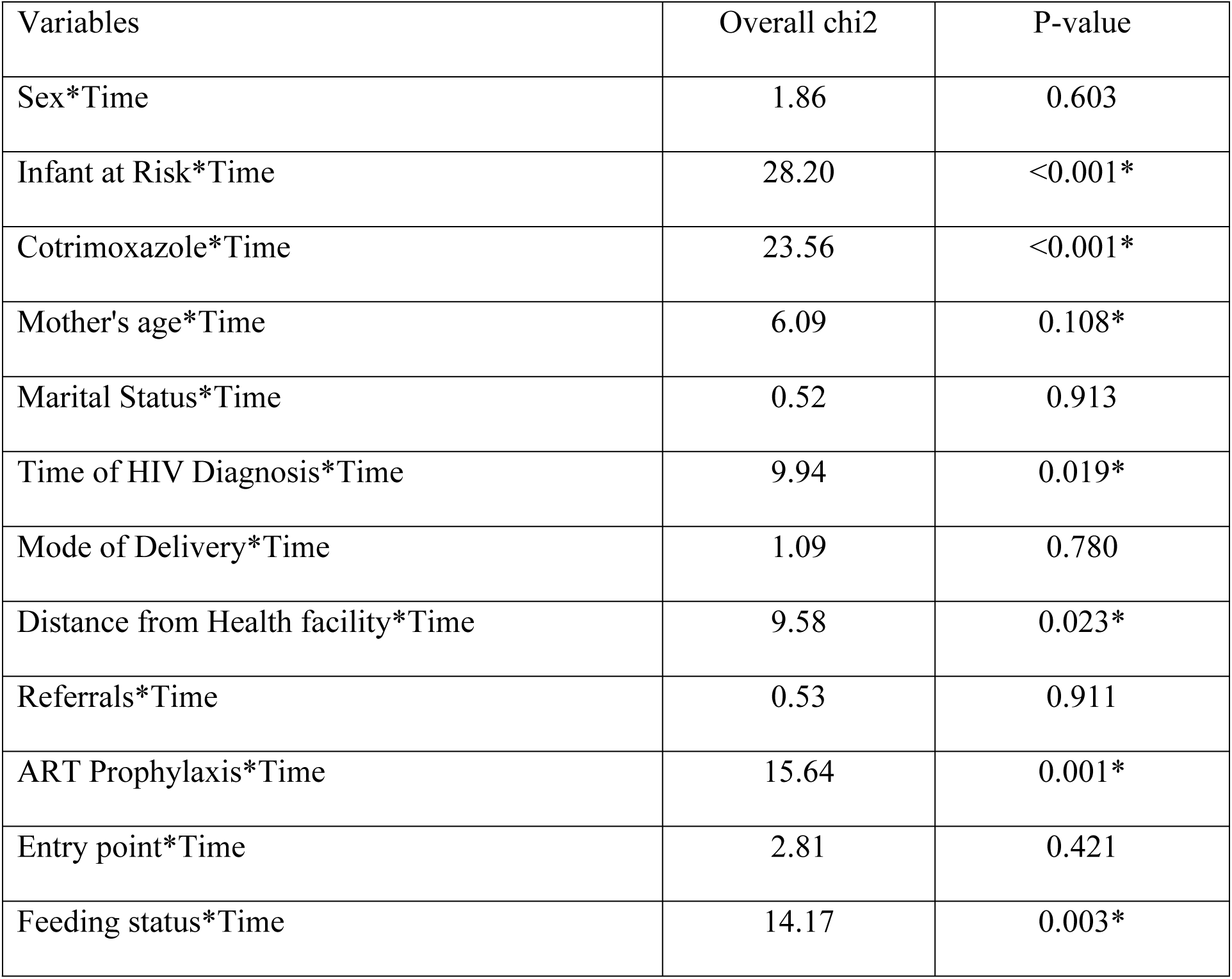
Bivariate analysis for variables associated with timely completion of EID overtime among 252 HIV exposed infants.

| Variables | Overall chi2 | P-value |
| --- | --- | --- |
| Sex*Time | 1.86 | 0.603 |
| Infant at Risk*Time | 28.20 | <0.001* |
| Cotrimoxazole*Time | 23.56 | <0.001* |
| Mother's age*Time | 6.09 | 0.108* |
| Marital Status*Time | 0.52 | 0.913 |
| Time of HIV Diagnosis*Time | 9.94 | 0.019* |
| Mode of Delivery*Time | 1.09 | 0.780 |
| Distance from Health facility*Time | 9.58 | 0.023* |
| Referrals*Time | 0.53 | 0.911 |
| ART Prophylaxis*Time | 15.64 | 0.001* |
| Entry point*Time | 2.81 | 0.421 |
| Feeding status*Time | 14.17 | 0.003* |

#### Multivariate analysis for the factors associated with timely completion of EID among HIV exposed infants at LRRH

Factors that were statistically associated with timely completion of EID included; No cotrimoxazole [aRR=2.97 95% CI (1.45-6.10)], categories of time, time 3 [aRR=6.45, 95% CI (4.50-9.25)], variable*Time interaction, No cotrimoxazole*Time2 [aRR=1.96, 95% CI (1.09-3.50)], No cotrimoxazole *Time3 [aRR=0.33, 95% CI (0.35-0.79)].

Results of multivariate analysis for the factors associated with timely completion of EID at LRRH are summarized in table 4.

**Table 4.** Multivariate analysis factors associated with timely completion of EID among 252 HIV exposed infants at LRRH.

| Variables | Category | aRR | 95% CI |  | p-values |
| --- | --- | --- | --- | --- | --- |
| Cotrimoxazole | Yes | Ref. |  |  |  |
|  | No | 2.97 | 1.45 | 6.10 | 0.003 |
| Time | 1 | Ref. |  |  |  |
|  | 2 | 0.79 | 0.49 | 1.27 | 0.323 |
|  | 3 | 6.45 | 4.50 | 9.25 | <0.001 |
|  | 4 | 1.26 | 0.81 | 1.97 | 0.300 |
| Cotrimoxazole*Time | No*1 | 1.75 | 0.68 | 4.48 | 0.244 |
|  | No*2 | 1.96 | 1.09 | 3.50 | 0.024 |
|  | No*3 | 0.32 | 0.35 | 0.79 | 0.013 |
|  | No*4 | 1.00 |  |  |  |

*aRR- Adjusted Relative Risk, 95% CI-95%Confidence Interval*

### Discussion

The overall uptake of EID among HEIs at 4-6 weeks for 1^st^ PCR, 9 months for 2^nd^PCR, 6 weeks after cessation of breastfeeding for 3^rd^PCR and 18 months for final RDT were 80.1%, 84.4%, 3.7% and 78.8% respectively. These rates were below the Ministry of Health (MOH) target of 90%.

The relatively higher uptake at 4-6 weeks (80.1%) and 9 months (84.4%) can be attributed to these time points coinciding with the immunization schedule. At 6 weeks, infants receive vaccinations for diphtheria, tetanus, pertussis, polio, and hepatitis B, while at 9 months, they receive measles-rubella I and yellow fever vaccines. This synchrony likely encourages mothers to bring their infants for both EID testing and immunization.

The extremely low uptake at 6 weeks after breastfeeding cessation (3.7%) could be due to incorrect appointment scheduling, with many mothers being instructed to return 2 months after breastfeeding cessation instead of 6 weeks, as per the guidelines.. Additionally, the third PCR test does not align with any immunization date, making it burdensome for mothers to return solely for EID testing, especially when most infants had tested HIV-negative by the 9-month mark.

At 18 months, the uptake of 78.8% may also be explained by the alignment with measles-rubella II vaccine, vitamin A supplementation, and deworming, which encourages mothers to bring their infants for the final HIV test. Mothers are also eager to have their infants discharged from the EID clinic at this point. However, the findings indicate that less than 90% of HEIs complete all EID tests, leaving a significant number of infants with unknown final HIV status. These findings are consistent with a study conducted among Ugandan fishing communities, where 81% of HEIs completed the final RDT, though only 6.1% completed the 2nd PCR within the recommended timeframe (Remegio et al., 2022). However, the uptake rates in this study were higher than those found in Kenya, where only 21.8% of HEIs completed the full series of 1st, 2nd, and 3rd PCR tests, and the final RDT (Mburu, 2021). This discrepancy could be due to differences in how “uptake” was defined.

Our study found that infants who did not receive cotrimoxazole prophylaxis were 2.43 times more likely to complete EID on time compared to those who received it. However, a retrospective study from Bishoftu Hospital in Ethiopia found no association between cotrimoxazole administration and EID uptake (12). The difference in findings could be attributed to the large sample size of 624 used compared to the small sample size of 252 used in this study.

There was a significant difference in timely completion of EID for the 3rd PCR, but not for other time points. This aligns with the There was a significant difference in timely completion of EID for the 3^rd^ PCR, but not for other time points. This aligns with the observation that the 3rd PCR does not coincide with any other scheduled healthcare activity, unlike other tests, which overlap with immunization visits.

ART prophylaxis was not significantly associated with timely completion of EID, (Kenya study, 2021). (Kenya HEI study, 2020). Our study found that ART prophylaxis was not statistically associated with timely completion of EID, consistent with findings from a study in Kenya, where HIV prophylaxis after delivery was not linked to poor health outcomes (13). However, a contrasting study among 2020 HEIs in Kenya showed that infants who did not receive ART prophylaxis were at higher risk for delayed EID (14). The larger sample size and multi-site nature of the Kenyan study likely provided more statistical power, accounting for the difference in results.

The findings from this study show that feeding status while plausible, but was not statistically associated with timely completion of EID. This finding aligns well with a study conducted among exposed infants in rural Uganda which also found no significant association between feeding status and EID uptake. This consistency could be attributed to the extensive research and promotion of breastfeeding among HIV positive mothers, which might mitigate the influence of feeding practices on EID. However, the findings from a cross sectional study on adherence to the HIV early infant diagnosis testing protocol among fishing communities of Uganda highlighted that cessation of breastfeeding was significantly associated with not receiving the 1^st^ DNA-PCR (11). The discrepancy could stem from methodological differences, as the fishing community study relied solely on quantitative methods, potentially overlooking broader contextual factors influencing EID completion. The discrepancy could be due to the methodological differences, as the fishing community study reied solely on only quantitative method, potentially overlooking broader contenxtual factors influencing EID completion.

The study found that distance from the health facility was another factor not statistically associated with timely completion of EID. This finding contrasts with the study conducted in Tanzania, where distance greater than 5 kilometers from health facilities was the most common reason for non-uptake of EID testing at six weeks after cessation of breastfeeding (15). Similarly, in a study conducted in rural southwestern Uganda, participants cited a lack of transport money as a barrier to utilizing EID services (16). Notably, the qualitative aspects of these studies uncovered the impact of distance on service uptake, indicating that mixed-method approaches might better capture such complexities of barriers.

The study found that mother’s age was found not to be statistically associated with timely completion of EID. This result can be explained by the fact that younger mothers, with an average age of 32 years, are often supported by their families, including spouses and guardians. The findings are consistent with a quasi-experimental study in Thyolo, Malawi, which reported that maternal age did not influence EID uptake at six weeks (17). However, our finding contradicts findings from a cross-sectional study from Kenya showing a significant association between caregivers’ age and poor health outcome with a p-value of 0.004 (13). The difference in findings may be due to differences in the mean age of the participants and study design. While this study utilized a retrospective cohort design, the Kenyan study employed a cross-sectional approach, which might better capture real-time variables impacting health outcomes. The study found that time of HIV diagnosis was not statistically associated with timely completion of EID. This could be attributed to intensive PMTCT services offered at ANC and during delivery. Supporting this, a retrospective cohort study in Myanmar identified maternal ART initiation before pregnancy as a risk factor for delayed or missed EID testing (18). Another cross-sectional study on factors associated with uptake of early infant diagnosis of HIV among HIV exposed infants in Mbeya, Tanzania found that HIV exposed infants born to mothers with known HIV status, were more likely to udergo timely EID(19). These differences may reflect variations in PMTCT service delivery and the influence of health system structures on EID adherence.

#### Limitations of the study

The quality of the records from which data was collected was suboptimal, as they were not designed for research purposes. Consequently, key variables such as maternal education level, place of delivery, number of children under five years in the household, household monthly income, whether the parent stays with the infant, and spouse support were not captured. These un-studied variables could have been potential confounders for the different relationship.

Selection bias due sampling bias introduced into the study due the kind of sampling method that was used. This then implies that our results are not generalizable since participants didn’t have some chance of being part of the study.

### Conclusion

This study revealed that the uptake of Early Infant Diagnosis (EID) among HIV-exposed infants at 4–6 weeks, 9 months, 6 weeks after cessation of breastfeeding, and 18 months was below the Ministry of Health (MoH) target of 90%. While timely administration of cotrimoxazole was associated with timely completion of EID, other factors such as infant risk status, maternal age, timing of HIV diagnosis, distance from health facilities, ART prophylaxis, and feeding status were not statistically significant. These findings underscore the need for targeted interventions to improve EID uptake and ensure timely testing of HIV-exposed infants to enhance health outcomes and support the achievement of MoH targets.

To enhance EID uptake, the MoH should strengthen supervision and mentorship to ensure adherence to guidelines while addressing gaps in service delivery. Future research should explore additional factors such as household income, parental education, and caregiver support, incorporating both rural and urban settings for broader comparisons. Improving record-keeping practices and engaging community health workers to raise awareness about EID schedules can further mitigate barriers. Tailored interventions, particularly for high-risk infants, and community-based education campaigns will be critical to achieving the MoH target and ensuring better health outcomes.

## Data Availability

All relevant data are within the manuscript and its Supporting Information files.

## List of abbreviations

AIDS: acquired immunodeficiency syndrome
ANC: Antenatal Care
ART: Antiretroviral Therapy
ARVs: Anti-retroviral drugs
DNA-PCR: Deoxyribonucleic acid-polymerase chain reaction
EID: Exposed Infant Diagnosis
EMTCT: Elimination of mother-to-child transmission
GEE: Generalized estimating equations
HEI: HIV Exposed Infant
HIV: Human Immunodeficiency Virus
HTS: HIV Testing Services
LRRH: Lira Regional Referral Hospital
MOH: Ministry of Health
MTCT: Mother-to-child transmission
PI: Principal Investigator
PMTCT: Prevention of mother-to-child transmission
RCT: Routine Counselling and testing
RDT: Rapid Diagnosis Test
SOMREC: School of Medicine Research Ethics Committee
UNAIDS: United Nations Programme on HIV/AIDS
UNICEF: United Nations International Children’s Emergency Fund
WHO: World Health Organization

## Declarations

### Conflicting Interests

The authors have no conflicting interests.

### Consent for publication

Not applicable.

### Availability of data materials

The datasets used and/or analyzed during the current study are available from the corresponding author on reasonable request.

### Competing interests

The authors declare no competing interests.

### Funding

The research received no specific grant from any funding agency in the public, commercial or not-for-profit sectors.

### Author’s contributions

Awili Rebecca: Contributed to study conceptualization, visualization, investigation, methodology, project administration, data collection and analysis.

Eve Nakabembe: Contributed to study conceptualization, methodology, supervision, validation, writing-review & editing.

Victor Musiime: Contributed to study conceptualization, methodology, supervision, validation, writing-review & editing.

Joan Kalyango: Contributed to study design, conceptualization, supervision, revision and conduct of the study.

Shaban Rajab Wilobo: Writing-review and editing

Nathan Ntenkaire: Writing-review and editing

Sean Steven Puleh: Contributed to validation, writing-review & editing.

Joy Acen: Contributed to validation, writing-review & editing.

Douglas Bulafu: Contributed to validation, writing-review & editing.

## Acknowledgements

We thank all the people who were involved in the synthesis of this research up to its completion, and grateful to Lira Regional Referral Hospital, research assistant, Supervisors, colleagues and lecturers at Clinical Epidemiology Unit, Makerere university.

## Supporting information

For objective 1; determine the uptake of EID

Using the formula for sample size calculations for longitudinal studies with a dichotomous outcome variable, estimates obtained from a cross sectional study done in the fishing area (11).

Assumed that subjects in the exposed and unexposed are equal (50%,50%)

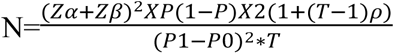

N is the required sample size

Z-alpha is the standard normal value corresponding to a 5% level of significance =1.96 Z-beta is the standard normal value corresponding with 80% power=0.84

P1-proportion of subjects with the outcome(cases) in the married group =0.411

P0-proportion of subjects with the outcome(cases) in the unmarried group group=0.241

P-is the weighted average of P1 and P0; 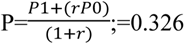

r-is the ratio in the number of subjects in the 2 groups (number in the married group=0.5 and number in the unmarried group=0.5); r=1

T-is the number of follow up measurements=3

ρ-is the correlation coefficient of the repeated measurements=0.25

N= 60

Adjusting for 10% missing data; N=66

## Notes

### Competing Interest Statement

The authors have declared no competing interest.

### Funding Statement

The author(s) received no specific funding for this work.

### Author Declarations

Makerere-School of Medicine Research Ethic Committee-2023-575

